# Association of Eicosanoids and Lung Function in MESA Lung and Framingham Heart Studies

**DOI:** 10.1101/2025.09.08.25335345

**Authors:** Mythri Ambatipudi, Jenna N. McNeill, Athar Roshandelpoor, Mona Alotaibi, Louisa A Mounsey, Eric A Hoffman, George O’Connor, Seung Hoan Choi, Norrina Allen, R Graham Barr, Mohit Jain, Susan Cheng, Jennifer E. Ho

**Author notes:** **Corresponding author:** Jennifer E. Ho, MD, Beth Israel Deaconess Medical Center 330 Brookline Avenue, E/CLS 945, Boston, MA 02215-5491.

## Abstract

**Introduction:** Eicosanoids are bioactive lipids with roles in airway remodeling, smooth muscle hypertrophy, emphysema and pulmonary fibrosis via inflammatory pathways. Specific eicosanoids have been associated with diseases like asthma and pulmonary fibrosis, yet their broader associations with lung function remain unclear. We investigated associations of eicosanoids and related metabolites with early changes in lung function and structure.

**Methods:** We comprehensively profiled >250 eicosanoids and eicosanoid-related metabolites using directed non-targeted mass spectrometry in the Multi-Ethnic Study of Atherosclerosis (MESA) Lung Study with independent validation in the Framingham Heart Study (FHS). We performed cross-sectional analysis of associations between metabolites and lung function as assessed by spirometry and quantitative computed tomography (CT) measures.

**Results:** Among 3384 MESA Lung participants (mean age 63±10 years, 51% women), 51 metabolites were associated with lung function (22 with % predicted FEV_1_, 18 with % predicted FVC, and 25 with FEV_1_/FVC), with 24 validated in FHS. Of these, 27 were associated with obstructive physiology, including linoleic acid derivatives (9-HODE) and other long-chain fatty acids (LCFAs, hydroxyhexadecanoic and hydroxyoctadecanoic acids) associated with higher odds. Fourteen metabolites were associated with restrictive physiology, including LTB3 and its analog associated with lower odds, and omega-3 fatty acids (EPA, stearidonic acid) associated with higher odds.

**Conclusions:** Specific eicosanoids and eicosanoid-related metabolites including linoleic acid derivatives and LCFAs were associated with obstructive, and leukotrienes and omega-3 fatty acids with restrictive physiology. These findings highlight bioactive lipids involved in pro- and anti-inflammatory pathways as potential influencers of lung function and may be future therapeutic targets.

**Take Home Message:** We identified eicosanoid metabolites associated with pulmonary function testing measurements and several that are associated with altered odds of obstructive and restrictive lung physiologies in a cohort MESA participants, with validation in FHS.

## INTRODUCTION

Chronic lung disease including chronic obstructive pulmonary disease (COPD), emphysema, pulmonary fibrosis, and asthma have impacts on health, quality of life, and mortality.(1–4) In 2017, chronic respiratory diseases affected 545 million people, accounting for 3.9 million deaths worldwide.(5)

While patient and population factors contribute to lung disease development, inflammation is a key pathway underlying both obstructive and restrictive diseases.(6) Eicosanoids are bioactive lipids derived from arachidonic acid (AA) that govern pro- and anti-inflammatory activity.(7) Prior studies have examined specific eicosanoid metabolites including leukotrienes and prostaglandins as mediators of various lung diseases in which they incite inflammation, regulate epithelial cell function, and promote fibrosis.(8–10) Additionally, leukotrienes contribute to airway remodeling via bronchial smooth muscle and fibroblast proliferation.(8) Targeting eicosanoid pathways has proven effective in lung disease via therapies including singulair (leukotrienes antagonist) and zileuton (5-lipoxygenase inhibitor) for asthma, and synthetic prostacyclins (iloprost) and others for pulmonary arterial hypertension.(11–13)

While specific eicosanoids have been implicated in lung disease, prior studies are limited to only dozens of metabolites, providing incomplete understanding of the broader bioactive lipid profile. We recently reproducibly ascertained >250 eicosanoid and related metabolites using a novel directed nontargeted liquid chromatography mass spectrometry (LC-MS)-based approach.(14) In this study, we leveraged this platform to investigate associations of eicosanoids and eicosanoid-related metabolites with early changes in lung function and structure across two community-based cohorts. By examining population-based samples, we examined the role of eicosanoids and eicosanoid-related metabolites as regulators of inflammation to determine their role in early disease and potentially inform therapeutic targets.

## METHODS

### Study Sample

The Multi-Ethnic Study of Atherosclerosis (MESA) recruited 6814 adults ages 45-84 who self-reported white, Black, Hispanic or Asian race/ethnicity and were free of clinical cardiovascular disease (CVD) from 2000-2002.(15) We included participants from examination cycle 2 (2002–2004) with plasma samples for eicosanoid analysis (n=5457). We excluded those with prevalent CVD (heart failure, myocardial infarction) (n=39), end-stage kidney disease (ESKD) (n=19), or missing clinical covariates (n=253), yielding 5150 participants. The MESA Lung Study performed spirometry in examination cycles 3-4 (2004-2007) among participants with genetic consent, endothelial function measures and with oversampling of Asian, 3489 of whom had eicosanoid measures;(16) 104 had incomplete/low-quality spirometry data, leaving 3385 participants. One participant was subsequently identified as a blood sample outlier, leaving 3384 participants **(Supplementary Figure 1)**.

We performed external validation among the Framingham Heart Study (FHS) Offpsring examination cycle 8 (2005–2008). Of the 2394 participants with plasma samples, we excluded those with prevalent CVD (n=32), ESKD (n=29), or missing clinical covariate data (n=11). Of the remaining 2321 patients, 2071 had complete spirometry measurements between 2005-2008.

All MESA and FHS participants gave written informed consent, and the study was approved by the appropriate institutional review boards.

### Clinical Assessment

Medical history, physical exam, and laboratory data were available for both MESA and FHS participants. Body mass index (BMI) was defined as weight/height^2^ (kg/m^2^) and ESKD as estimated glomerular filtration rate (eGFR) < 30 ml/min/1.73 m^2^. Smoking status was self-reported.

### Pulmonary Function Testing

MESA Lung participants underwent pre-bronchodilator spirometry using rolling barrel spirometers (OMI systems) per ATS/ERS guidelines.(17) Spirometry and diffusing capacity of the lungs for carbon monoxide (DLCO) measurements were collected for FHS using the Collins Comprehensive Pulmonary Laboratory system (nSpire Health Inc., Longmot, CO, USA).(18) Percent predicted FEV_1_ and FVC (PPFEV_1_, PPFVC) were calculated using published reference values and equations.(19,20) As per prior manuscripts, we utilized predicted spirometry values based on Hankinson equations. Restrictive physiology was defined as FEV_1_/FVC>0.7 and PPFVC<80%.(21) Participants with FEV_1_/FVC<0.7 and 80%<PPFEV_1_<100% were classified as Global Initiative for Obstructive Lung Disease (GOLD) grade 1 obstructive physiology while those with FEV_1_/FVC<0.7 and PPFEV_1_<80% had GOLD grade 2-4 obstructive physiology (combined into one due to small sample sizes in grades 3-4).(21–23)

### Lung Imaging

MESA participants underwent low-dose computed tomography (CT) for coronary artery calcium at exam 1 (2000–2002).(24) Images were analyzed for high attenuation areas (HAA), defined as percent lung volume with CT attenuation between -600 and -250 Hounsfield Units (HU),(25) and percent emphysema defined as percent of voxels <-950 HU.(25) Exam 5 (2010-2012) included full-lung CT scans,(26) with interstitial lung abnormalities (ILA) defined as ground-glass, reticular abnormality, diffuse centrilobular nodularity, honeycombing, traction bronchiectasis, non-emphysematous cysts or architectural distortion in ≥5% of nondependent lung regions.(27,28) Continuous HAA was created through log2-transformation of percent HAA.(25) ILA was analyzed binarily as absence of ILA versus indeterminate/definitive ILA.(28)

### Plasma Metabolite Profiling

Eicosanoid profiling for MESA and FHS was performed at UC San Diego. Plasma samples were drawn after ≥8 hours fasting and stored at -80°C. Eicosanoids and related metabolites were assayed via directed, non-targeted LC-MS.(14) Commercially-available standards and MS/MS fragmentation patterns were used to annotate metabolites, which were validated using spectral fragmentation pattern networking and manual annotation.

Due to informative missingness indicating concentrations below detectable threshold, missing values were imputed as 25% of the minimum value of that metabolite across participants. Low-abundance metabolites with >90% of data missing across participants were excluded. In total, 784 eicosanoids and related metabolites (98% of total) were included in subsequent MESA analyses. Metabolite alignment between MESA and FHS was determined via peak matching using m/z (mass-to-charge ratios) and retention times. In total, 454 metabolites were present in both MESA and FHS and considered for external validation.

### Statistical Analysis

Baseline characteristics were summarized with continuous variables as mean (SD) or median (IQR) and categorical variables as number (percentage). Metabolite concentrations were natural log-transformed for right skewness and standardized to mean 0 and standard deviation (SD) 1. In principal component analysis, an outlier was identified and excluded. In primary analyses, we investigated associations between metabolites and lung function measured by PPFEV_1_, PPFVC, and FEV_1_/FVC using multivariable linear regression models. Models were adjusted for age, sex, plate number, race, BMI, diabetes, hypertension, and aspirin/statin use.

Secondary models additionally adjusted for smoking status. False discovery rate (FDR)-adjusted q<0.05 was deemed significant for both MESA and FHS. Significant metabolites were tested for DLCO associations in FHS (unavailable in MESA).

In secondary analyses, we examined associations of metabolites with obstructive and restrictive physiology, with individuals who did not meet obstructive or restrictive criteria as the comparator group and FDR-adjusted q<0.1 deemed signficant. Logistic regression models were adjusted for the same covariates as in primary analyses. We used FDR-adjusted q<0.1 to validate results in FHS. Among the 2233 MESA participants with lung imaging, we examined associations of spirometry-associated metabolites with percent HAA, presence of ILA, and percent emphysema. Multivariable regression models were adjusted for covariates listed above, plus CT scanner type and wt220 (whether weight>220 lbs in Exams 1-4, MDCT scanners only). P<0.05 was deemed signficant.

In exploratory analyses, we tested eicosanoids as mediators of the interaction between smoking status and obstructive physiology. Mediation analysis was performed using the “mediation” package in R, with eicosanoids as mediators, obstructive physiology as the outcome, and smoking status (never versus past/current) as the exposure. Magnitudes and directionalities of exposure-mediator, mediator-outcome, and exposure-outcome associations were examined, along with sizes of mediation effects. Significant mediation was defined as mediation p<0.05.

All analyses were performed in RStudio (R version 4.2.1).

## RESULTS

Among 3384 MESA participants (mean age 63±10, 51% female), 44% had hypertension, 13% diabetes, and 11% were current smokers, with mean BMI 28.1±5.3 kg/m^2^. Most had normal lung physiology with median PPFEV_1_ of 100% (Q1 92%, Q3 110%), PPFVC of 98% (Q1 90%, Q3 108%), and FEV_1_/FVC of 0.78 (Q1 0.74, Q3 0.81) (**Table 1**). Overall, 989 (29.2%) had abnormal physiology, with 18.6% obstructive (8.7% grade 1, 9.9% grade 2-4) and 10.7% restrictive physiology.

**Table 1:** Clinical characteristics of MESA and FHS participants.

|  | MESA |  |  |  | FHS |  |  |  |
| --- | --- | --- | --- | --- | --- | --- | --- | --- |
|  | Normal<br>N = 2395 | Restriction<br>N = 361 | Grade 1<br>Obstruction<br>(N = 294) | Grade 2-4<br>Obstruction<br>(N = 334) | Normal<br>N = 1467 | Restriction<br>N = 94 | Grade 1<br>Obstruction<br>N = 273 | Grade 2-4<br>Obstruction<br>N = 237 |
| Clinical Measures |  |  |  |  |  |  |  |  |
| Age, years | 62 (10) | 63 (9) | 67 (9) | 66 (9) | 66 (9) | 65 (9) | 67 (9) | 69 (8) |
| Female, N (%) | 1272 (53) | 206 (57) | 99 (34) | 138 (41) | 853 (58) | 50 (53) | 132 (48) | 141 (60) |
| Current smoker, N (%) | 190 (8) | 44 (12) | 49 (17) | 71 (21) | 75 (5) | 16 (17) | 33 (12) | 47 (20) |
| Former smoker, N (%) | 875 (37) | 112 (31) | 133 (45) | 168 (50) | 455 (31) | 23 (25) | 116 (43) | 117 (49) |
| Pack years smoking, years | 9 (17) | 14 (29) | 21 (28) | 27 (31) | 11 (17) | 16 (22) | 22 (24) | 33 (27) |
| BMI, kg/m<sup>2</sup> | 28 (5) | 30 (6) | 27 (4) | 28 (5) | 28 (5) | 32 (7) | 28 (5) | 28 (6) |
| Hypertension, N (%) | 978 (41) | 187 (52) | 139 (47) | 175 (52) | 977 (67) | 79 (84) | 195 (71) | 175 (74) |
| Diabetes mellitus, N (%) | 288 (12) | 81 (22) | 25 (9) | 50 (15) | 166 (11) | 27 (29) | 32 (12) | 40 (17) |
| Spirometry Measures, median (Q1, Q3) |  |  |  |  |  |  |  |  |
| FEV1, % predicted | 100 (92, 110) | 76 (68, 81) | 89 (84, 94) | 69 (58, 76) | 103 (95, 112) | 75 (70, 80) | 90 (85, 94) | 69 (62, 76) |
| FVC, % predicted, | 98 (90, 108) | 73 (68, 77) | 102 (96, 108) | 84 (75, 91) | 104 (95, 114) | 74 (70, 77) | 103 (99, 108) | 86 (79, 94) |
| FEV1/FVC ratio | 0.78 (0.74, 0.81) | 0.79 (0.76, 0.83) | 0.67 (0.63, 0.69) | 0.63 (0.55, 0.67) | 0.75 (0.72, 0.78) | 0.77 (0.73, 0.80) | 0.66 (0.63, 0.68) | 0.61 (0.55, 0.65) |
| DLCO<sup>1</sup> |  |  |  |  | 22 (18, 27) | 19 (15, 22) | 21 (18, 26) | 18 (15, 22) |
| CT measures (n=2233)<sup>2</sup> |  |  |  |  |  |  |  |  |
| % Emphysema | 4 (4) | 2 (2) | 6 (4) | 6 (5) |  |  |  |  |
| % High attenuation areas | 7 (4) | 9 (7) | 6 (3) | 6 (2) |  |  |  |  |
| Interstitial lung abnormalities, N (%) | 401 (25) | 52 (24) | 69 (39) | 64 (31) |  |  |  |  |

Values are mean (SD) unless specified.
<sup>1</sup> 1972 FHS participants had DLCO measures (Normal: 1441; Restrictive: 87; Obstructive: 259 grade 1, 273 grade 2-4)
<sup>2</sup> 2233 MESA participants had CT measures (Normal: 1630; Restrictive: 216 restrictive; Obstructive: 178 grade 1, 209 grade 2-4)

### Associations of Eicosanoids with Spirometry Variables in MESA

Of the 784 eicosanoids or related metabolites, 51 were associated with at least one spirometry measure in MESA (FDR q<0.05, **Supplementary Table 1, Figure 1),** 19 with known molecular identities (**Table 2)**. Metabolites associated with higher PPFEV_1_ included a leukotriene B3 (LTB3) analog and docosahexaenoic acid (DHA)-derivative maresin 1 **(Figure 2a)**. By contrast, linoleic acid (LA)-derivative 9-hydroxyoctadecadienoic acid (9-HODE), dihomo-gamma linolenic acid (DGLA)-derivative hydroxy-eicosatrienoic acid (HETrE), and long-chain fatty acids (LCFAs) including palmitic and stearic acid derivatives (hydroxyhexadecanoic acid, hydroxyoctadecanoic acids) correlated with lower PPFEV_1_ **(Table 2)**.

**Figure 1:**
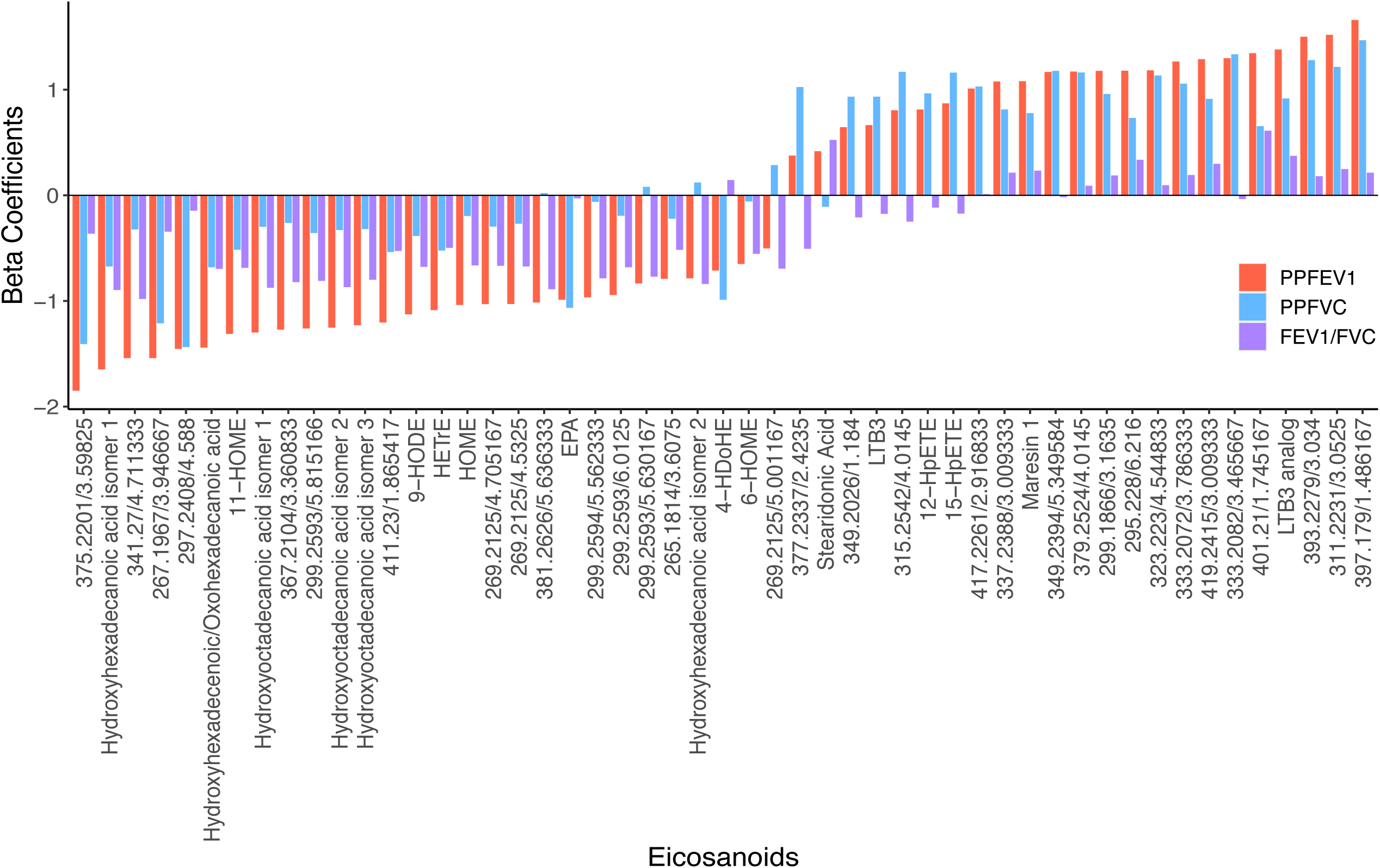
Association of eicosanoids with lung function, including PPFEV1 (red), PPFVC (blue), and FEV1/FVC (purple). 51 metabolites displayed at least one significant association with spirometry traits. Beta estimates represent multivariable-adjusted associations between eicosanoids and spirometry variables. Metabolites are designated with putative ID if known and mass-to-charge ratio/retention time (min) if not.

**Figure 2:**
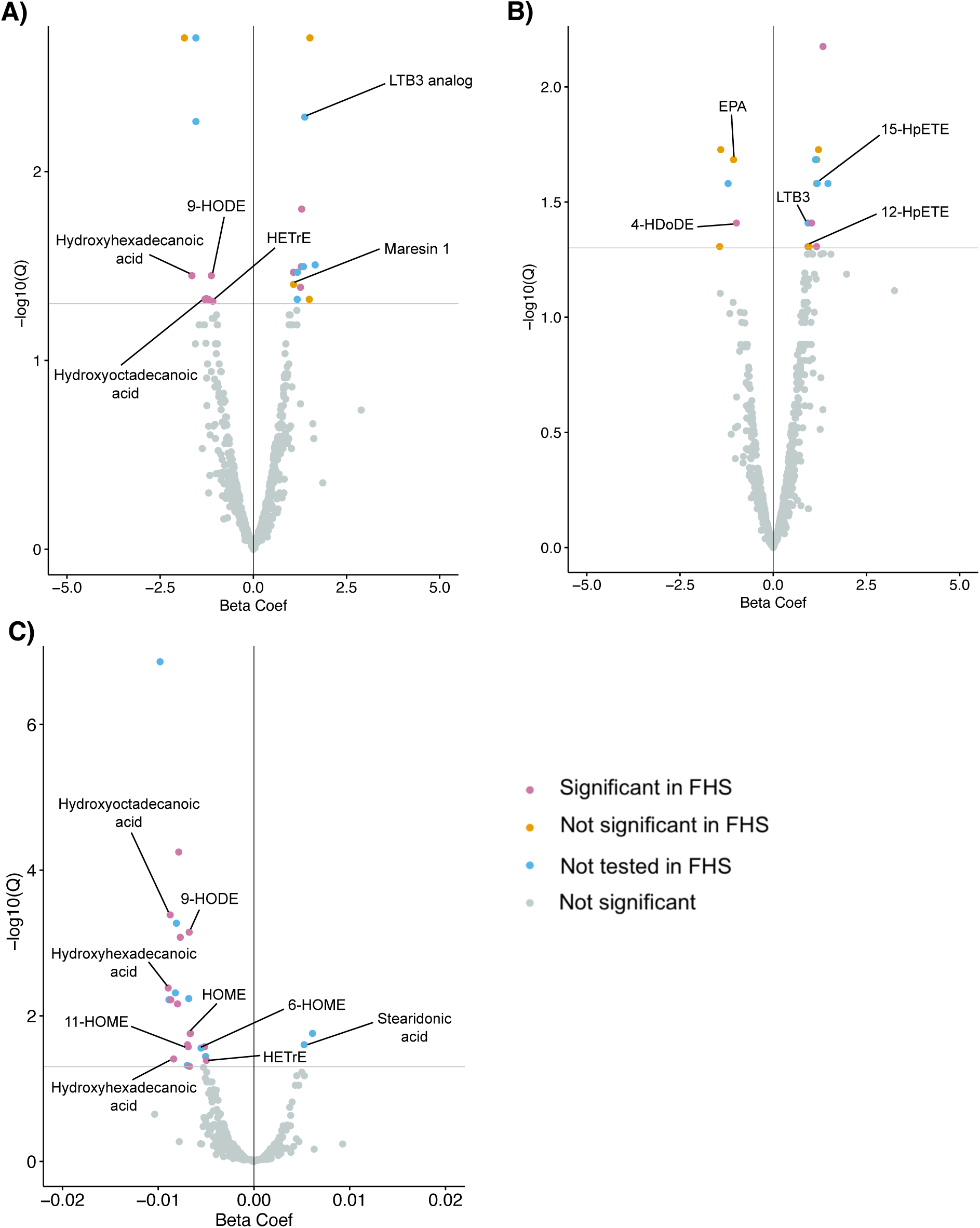
Associations of eicosanoids with A) PPFEV1, B) PPFVC, and C) FEV1/FVC. Beta coefficients indicate strength of associations. Grey line indicates q<0.05 with n=22 significant for PPFEV1, n=18 for PPFVC, and n=25 for FEV1/FVC. Coloring indicates those available for validation that did (pink) and did not (orange) validate in FHS.

**Table 2:**
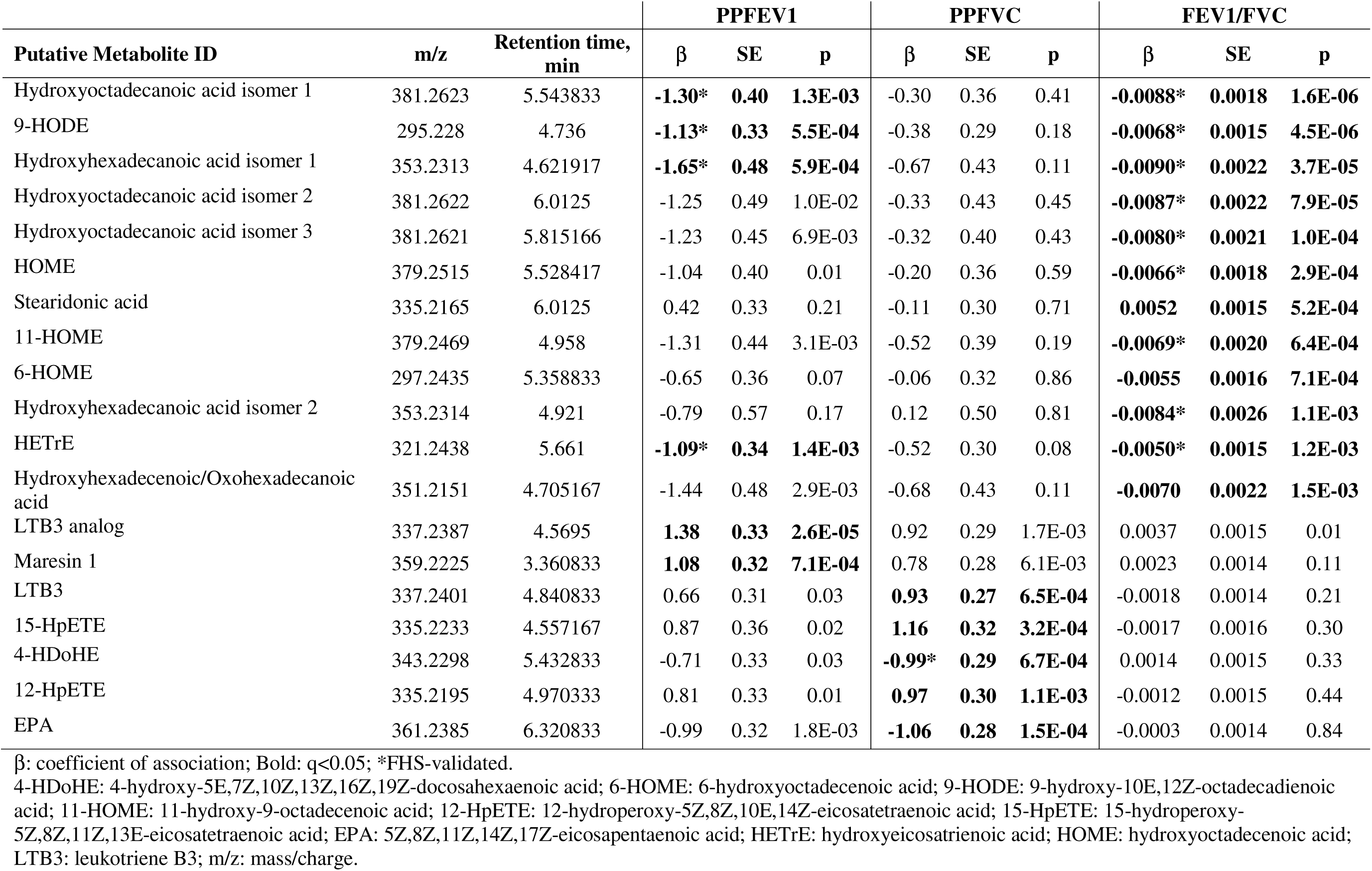
Associations between eicosanoids and spirometry measurements for metabolites with known identities.

Eighteen metabolites were associated with PPFVC **(Figure 2b)**. AA-derivatives 15-hydroperoxyeicosatetraenoic acid and 12-hydroperoxyeicosatetraenoic (15-HpETE, 12-HpETE), along with LTB3 correlated with higher PPFVC, while eicosapentaenoic acid (EPA) and DHA-derivative 4-hydroxydocosahexaenoic acid (4-HDoHE) correlated with lower PPFVC **(Table 2)**.

Twenty-five metabolites were associated with FEV_1_/FVC ratio **(Figure 2c)**. Stearidonic acid correlated with higher FEV_1_/FVC. Palmitic acid derivatives (hydroxyhexadecanoic acid, oxohexadecanoic acid), stearic acid-derivative hydroxyoctadecanoic acid, DGLA-derivative HETrE, and LA derivatives (9-HODE, 6-hydroxyoctadecadienoic acid (6-HOME), 11-hydroxyoctadecenoic acid (11-HOME)) correlated with lower FEV_1_/FVC **(Table 2)**.

### Validation of Spirometry-Associated Eicosanoids in FHS

Among 22 metabolites associated with PPFEV_1_ in MESA, 13 were available for validation in FHS, with 9 validating **(Figure 2a)**. Directionality was consistent for all PPFEV_1_-associated metabolites that validated (**Figure 3a**), which included 9-HODE, hydroxyhexadecanoic acid, hydroxyoctadecanoic acid, and HETrE, all associated with lower PPFEV_1_ across both cohorts.

**Figure 3:**
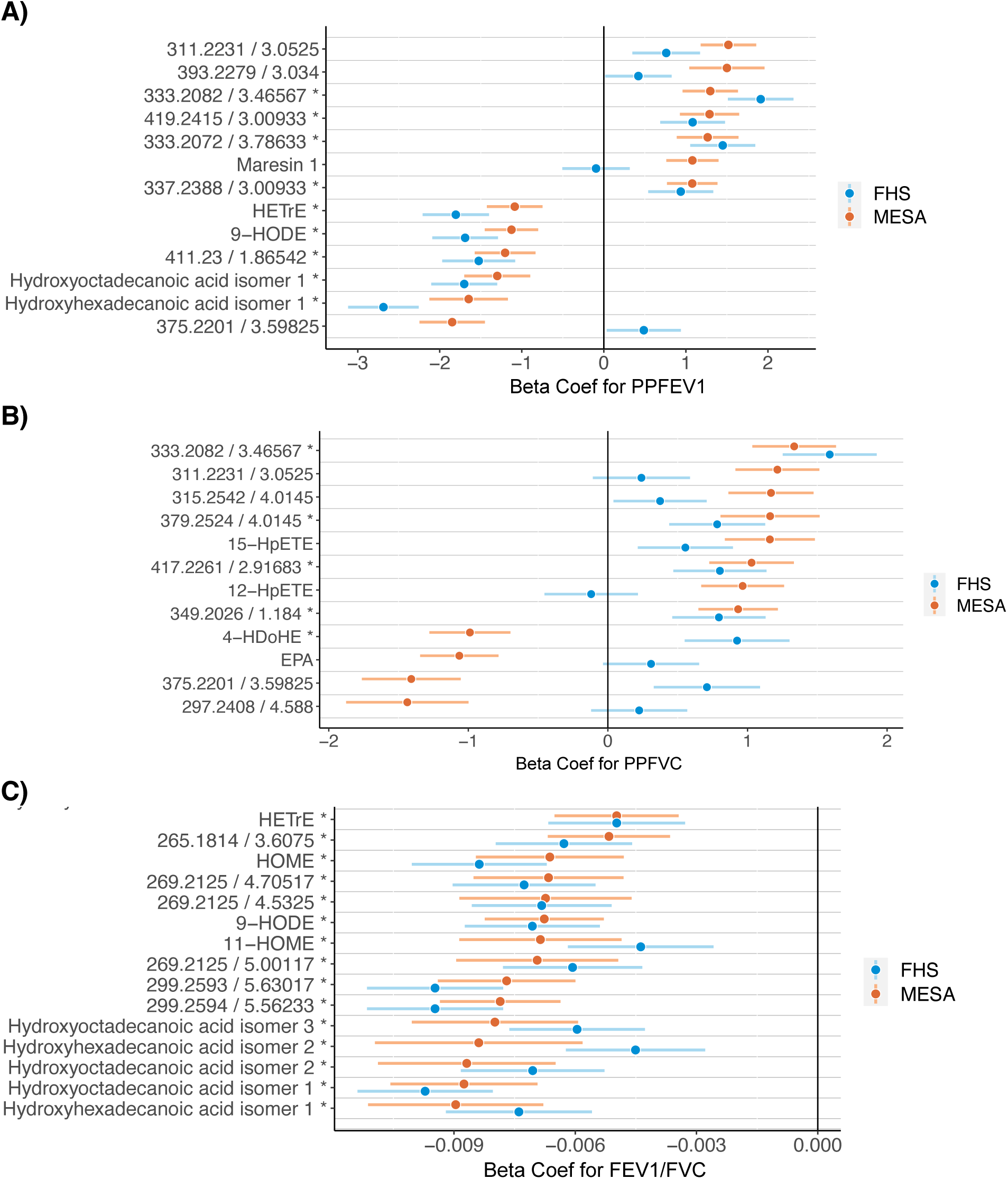
Eicosanoids significantly associated with A) PPFEV1, B) PPFVC, and C) FEV1/FVC in MESA (orange) and available for validation in FHS (blue). Beta estimates represent strength of associations. Metabolites are designated with putative ID if known and mass-to-charge ratio/retention time (min) if not. *Validated in FHS participants.

Among 18 metabolites associated with PPFVC in MESA, 12 were available in FHS and 5 validated **(Figure 2b)**. Validated metabolites were directionally concordant, excepting DHA-derivative 4-HDoHE which correlated with lower PPFVC in MESA and higher PPFVC in FHS **(Figure 3)**.

Lastly, among 25 metabolites associated with FEV_1_/FVC in MESA, 15 were available in FHS, and all validated, with all metabolites associated with lower FEV_1_/FVC across both cohorts (**Figure 3c**).

We examined associations with DLCO in FHS. Among 51 metabolites associated with at least one spirometry measure in MESA, 33 were available in FHS, with 18 associated with DLCO. Thirteen metabolites correlated with lower DLCO and directionally consistent across other phenotypes, including LA-derivatives 9-HODE and 11-HOME and palmitic and stearic acid-derivatives hydroxyhexadecanoic and hydroxyoctadecanoic acids, which correlated with lower PPFEV_1_ and FEV_1_/FVC, as well as AA-derivative 12-HpETE which correlated with greater PPFVC. **(Supplementary Table 1)**.

### Associations of Significant Eicosanoids with Obstructive and Restrictive Physiology

Of the 51 metabolites associated with spirometry measures, 27 were associated with obstructive and 14 with restrictive physiology in MESA **(Table 3A-3B)**.

**Table 3:** Associations between eicosanoids and obstructive and restrictive lung disease.

| Putative Metabolite ID | m/z | Retention time, min | OR | 95% CI | P |
| --- | --- | --- | --- | --- | --- |
| **A. Obstructive Disease** |  |  |  |  |  |
| *Associated with higher odds* |  |  |  |  |  |
| Unknown | 341.27 | 4.711333 | 1.35 | (1.25, 1.44) | 2.0E-09 |
| Unknown | 375.2201 | 3.59825 | 1.25 | (1.12, 1.38) | 6.3E-04 |
| Hydroxyhexadecanoic acid isomer 1* | 353.2313 | 4.621917 | 1.25 | (1.11, 1.40) | 2.4E-03 |
| Unknown | 367.2104 | 3.360833 | 1.24 | (1.08, 1.40) | 8.9E-03 |
| Hydroxyhexadecenoic/Oxo-hexadecanoic acid | 351.2151 | 4.705167 | 1.22 | (1.07, 1.37) | 8.9E-03 |
| Unknown | 299.2593 | 5.815166 | 1.21 | (1.10, 1.33) | 8.7E-04 |
| Hydroxyoctadecanoic acid isomer 1* | 381.2623 | 5.543833 | 1.21 | (1.10, 1.33) | 9.9E-04 |
| Unknown* | 269.2125 | 4.705167 | 1.21 | (1.09, 1.34) | 2.2E-03 |
| Unknown* | 269.2125 | 4.5325 | 1.21 | (1.07, 1.35) | 8.6E-03 |
| Hydroxyoctadecanoic acid isomer 2* | 381.2622 | 6.0125 | 1.21 | (1.06, 1.36) | 0.01 |
| Unknown* | 299.2594 | 5.562333 | 1.19 | (1.10, 1.28) | 1.8E-04 |
| Unknown* | 299.2593 | 5.630167 | 1.19 | (1.08, 1.30) | 1.5E-03 |
| Unknown* | 265.1814 | 3.6075 | 1.19 | (1.08, 1.30) | 1.6E-03 |
| Unknown | 381.2626 | 5.636333 | 1.19 | (1.04, 1.34) | 0.02 |
| Hydroxyoctadecanoic acid isomer 3* | 381.2621 | 5.815166 | 1.18 | (1.04, 1.32) | 0.02 |
| 9-HODE* | 295.228 | 4.736 | 1.17 | (1.08, 1.27) | 1.1E-03 |
| Unknown | 299.2593 | 6.0125 | 1.17 | (1.05, 1.28) | 7.4E-03 |
| Unknown* | 269.2125 | 5.001167 | 1.17 | (1.04, 1.31) | 0.02 |
| Unknown | 267.1967 | 3.946667 | 1.15 | (1.04, 1.27) | 0.01 |
| 6-HOME | 297.2435 | 5.358833 | 1.14 | (1.03, 1.26) | 0.02 |
| Unknown* | 411.23 | 1.865417 | 1.14 | (1.02, 1.25) | 0.03 |
| *Associated with lower odds* |  |  |  |  |  |
| LTB3 analog | 337.2387 | 4.5695 | 0.89 | (0.79, 0.99) | 0.02 |
| Stearidonic Acid | 335.2165 | 6.0125 | 0.88 | (0.78, 0.98) | 0.01 |
| Maresin 1 | 359.2225 | 3.360833 | 0.88 | (0.78, 0.99) | 0.02 |
| Unknown | 295.228 | 6.216 | 0.86 | (0.75, 0.98) | 0.01 |
| Unknown | 397.179 | 1.486167 | 0.86 | (0.72, 1.00) | 0.03 |
| Unknown | 401.21 | 1.745167 | 0.84 | (0.73, 0.96) | 3.5E-03 |
| **B. Restrictive Disease** |  |  |  |  |  |
| *Associated with higher odds* |  |  |  |  |  |
| EPA | 361.2385 | 6.320833 | 1.21 | (1.09, 1.34) | 3.0E-03 |
| Stearidonic Acid | 335.2165 | 6.0125 | 1.20 | (1.06, 1.35) | 0.01 |
| Unknown | 267.1967 | 3.946667 | 1.19 | (1.06, 1.33) | 9.4E-03 |
| *Associated with lower odds* |  |  |  |  |  |
| Unknown* | 417.2261 | 2.916833 | 0.86 | (0.73, 0.99) | 0.02 |
| Unknown | 299.1866 | 3.1635 | 0.85 | (0.72, 0.97) | 0.01 |
| Unknown | 311.2231 | 3.0525 | 0.85 | (0.71, 0.99) | 0.02 |
| Unknown | 323.223 | 4.544833 | 0.85 | (0.72, 0.98) | 0.02 |
| Unknown | 333.2072 | 3.786333 | 0.84 | (0.70, 0.99) | 0.02 |
| LTB3 | 337.2401 | 4.840833 | 0.83 | (0.69, 0.97) | 7.4E-03 |
| LTB3 analog | 337.2387 | 4.5695 | 0.83 | (0.69, 0.97) | 7.9E-03 |
| Unknown | 377.2337 | 2.4235 | 0.82 | (0.68, 0.95) | 2.9E-03 |
| Unknown | 349.2394 | 5.349584 | 0.82 | (0.67, 0.96) | 5.6E-03 |
| Unknown* | 333.2082 | 3.465667 | 0.81 | (0.68, 0.93) | 5.9E-04 |
| Unknown* | 393.2279 | 3.034 | 0.75 | (0.55, 0.94) | 3.0E-03 |

Significance defined as FDR $q < 0.1$ . Putative metabolite IDs shown for metabolites with known confirmed identities.
\*Validated in FHS participants.
6-HOME: 6-hydroxyoctadecenoic acid; 9-HODE: 9-hydroxy-10E,12Z-octadecadienoic acid; EPA: 5Z,8Z,11Z,14Z,17Z-eicosapentaenoic acid; LTB3: leukotriene B3; m/z: mass/charge; OR: odds ratio.

Of the 27 metabolites associated with obstructive physiology, 14 were available in FHS, and 12 replicated. Metabolites with significant associations in both MESA and FHS included 9-HODE, a hydroxyhexadecanoic acid isomer, and hydroxyoctadecanoic acid isomers, all of which were associated with greater odds of obstructive physiology. Specifically, a 1-SD higher hydroxyoctadecanoic acid isomer 1 was associated with 1.21-fold (95% CI 1.10-1.33) greater odds of obstruction. Similarly, a 1-SD higher 9-HODE was associated with 1.17-fold (95% CI 1.08-1.27) higher odds of obstruction **(Figure 4A)**. Most metabolites with significant associations in only MESA participants were also associated with greater odds of obstruction, including stearidonic acid and 6-HOME. Only 6 metabolites were associated with lower odds of obstructive physiology, including an LTB3 analog and maresin 1 **(Supplementary Figure 3)**.

**Figure 4:**
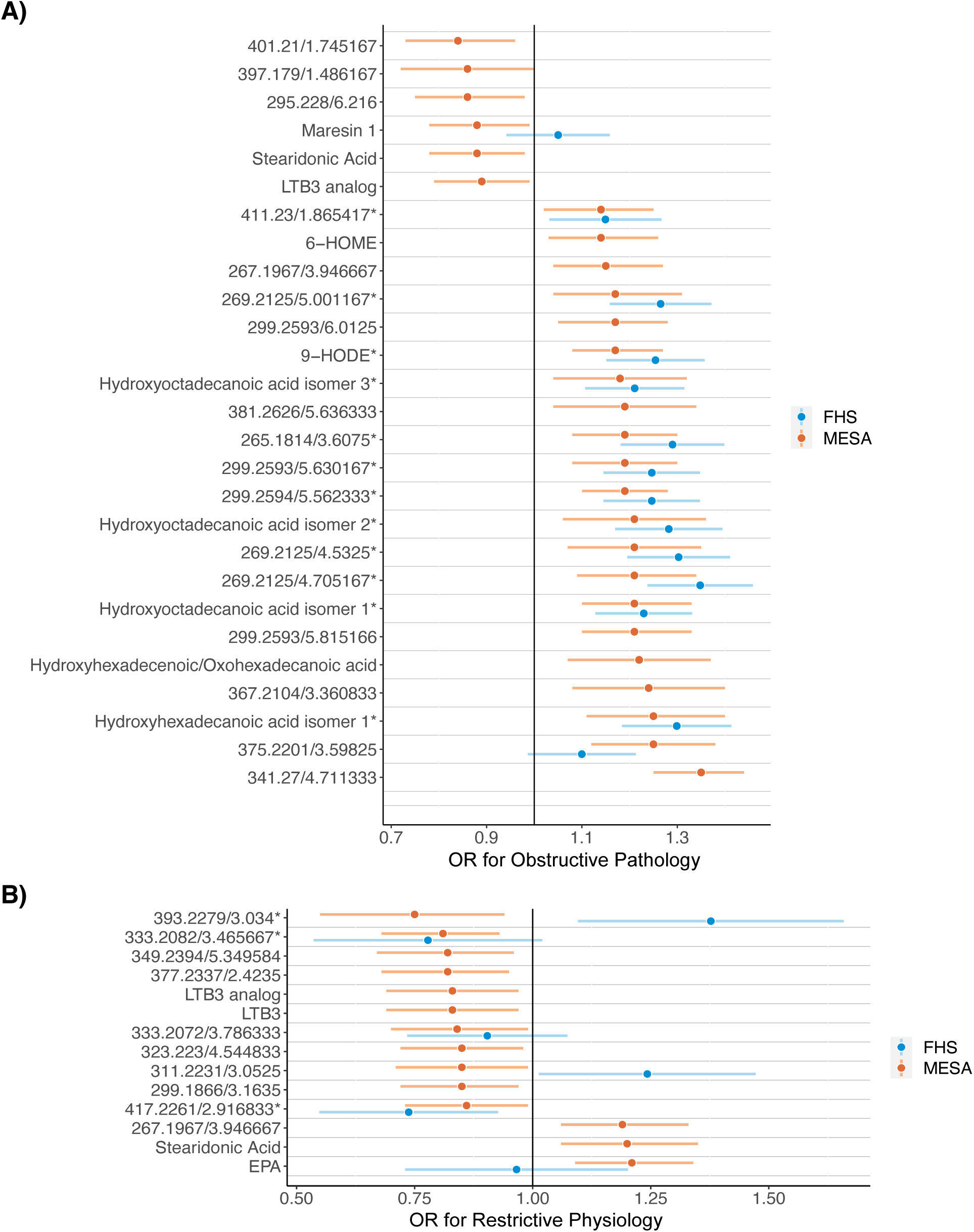
Associations of eicosanoids with A) obstructive or B) restrictive physiology in MESA (orange) and FHS (blue). Odds ratios represent odds of obstructive vs normal, or restrictive vs normal, per 1-SD higher log-transformed metabolite. Metabolites are designated with putative ID if known and mass-to-charge/retention time (min) if not. *Validated in FHS

Of the 14 metabolites associated with restrictive physiology in MESA, 6 were available in FHS, and 3 replicated. Although molecular identities of these 3 metabolites were unknown, all 3 were associated with lower odds of restrictive physiology. Most of the metabolites associated with restrictive physiology in only MESA participants were also associated with lower odds, including LTB3 and its analog (OR 0.83 [0.69-0.97]). By contrast, stearidonic acid and EPA were associated with greater odds of restrictive physiology (OR 1.20 [1.06-1.35]; OR 1.21 [1.09-1.34], respectively; **Figure 4B)**.

### Smoking and Association of Eicosanoids with Obstructive Physiology

Of the 3384 MESA participants, 354 (10.5%) were current and 1288 (38.1%) former smokers. Twenty-six of 51 metabolites from primary analyses remained significantly associated with at least one spirometry variable after additionally adjusting for smoking status **(Supplementary Table 2)**, with similar effect sizes and directionality **(Supplementary Figure 2)**.

Mediation analysis showed that of the 27 metabolites associated with obstructive physiology in MESA, smoking status mediated the association of 5 metabolites with obstructive physiology **(Supplementary Table 3)**. This included 9-HODE (0.90% effect mediated), maresin 1 (3.3%), and a hydroxyoctadecanoic acid isomer (2.6%). Most metabolites demonstrated mediation effects of small magnitudes, with the largest being approximately 4%.

### Associations of Significant Eicosanoids with Lung Imaging Features

Among 2233 MESA participants with CT scans, mean % emphysema was 4.1 ± 3.9, mean % HLA 6.8 ± 4.5, and 586 (22.6%) had ILA. Percent emphysema was in obstructive physiology (grade 1: 6.1±4.4, grade 2-4: 6.1± 5.4) versus normal/restrictive physiology (3.8±3.7; 2.3±2.3). Similarly, % HAA was in restrictive physiology (8.9±6.5) versus obstructive physiology (grade 1: 5.7±2.9, grade 2-4: 5.8±2.2) (**Table 1**).

Of the 51 metabolites associated with spirometry measures in MESA, 5 were associated with % emphysema, including LTB3 analog (previously associated with higher PPFEV_1_) which correlated with lower % emphysema (**Table 4A**). Six metabolites were associated with % HAA (**Table 4A-4C**), including HETrE, LTB3, and LTB3 analog, which correlated with greater % HAA, and 11-HOME which correlated with lower % HAA. Three metabolites were associated with ILA yet did not have confirmed known identities.

**Table 4:** Associations between eicosanoids and lung imaging features.

| **A: Percent Emphysema** |  |  |  |  |  |
| --- | --- | --- | --- | --- | --- |
| **Putative Metabolite ID** | **m/z** | **Retention time, min** | **$\beta$** | **SE** | **P** |
| Unknown | 341.27 | 4.711333 | -0.29 | 0.09 | 7.5E-04 |
| Unknown | 411.23 | 1.865417 | 0.26 | 0.09 | 0.00 |
| Unknown | 299.1866 | 3.1635 | 0.23 | 0.09 | 0.01 |
| Unknown | 401.21 | 1.745167 | -0.22 | 0.09 | 0.01 |
| LTB3 analog | 337.2387 | 4.5695 | -0.18 | 0.08 | 0.04 |
| **B: HAA Continuous** |  |  |  |  |  |
| **Putative Metabolite ID** | **m/z** | **Retention time, min** | **$\beta$** | **SE** | **P** |
| Unknown | 401.21 | 1.745167 | 0.04 | 0.01 | 1.3E-03 |
| HETrE | 321.2438 | 5.661 | 0.03 | 0.01 | 3.4E-02 |
| LTB3 | 337.2401 | 4.840833 | 0.03 | 0.01 | 0.02 |
| LTB3 analog | 337.2387 | 4.5695 | 0.03 | 0.01 | 0.02 |
| Unknown | 341.27 | 4.711333 | 0.03 | 0.01 | 0.05 |
| 11-HOME | 379.2469 | 4.958 | -0.04 | 0.02 | 0.04 |
| **C: HAA Dichotomized** |  |  |  |  |  |
| **Putative Metabolite ID** | **m/z** | **Retention time, min** | **OR** | **95% CI** | **P** |
| Unknown | 299.1866 | 3.1635 | 0.81 | (0.68, 0.94) | 0.00 |
| Unknown | 299.2593 | 6.0125 | 0.83 | (0.69, 0.97) | 0.01 |
| 11-HOME | 379.2469 | 4.958 | 0.80 | (0.63, 0.97) | 0.01 |
| Hydroxyoctadecanoic acid isomer 2 | 381.2622 | 6.0125 | 0.78 | (0.59, 0.96) | 0.01 |
| Unknown | 401.21 | 1.745167 | 1.17 | (1.04, 1.31) | 0.02 |
| LTB3 | 337.2401 | 4.840833 | 1.13 | (1.03, 1.24) | 0.02 |
| Unknown | 311.2231 | 3.0525 | 0.87 | (0.75, 0.99) | 0.03 |
| 6-HOME | 297.2435 | 5.358833 | 0.87 | (0.74, 1.01) | 0.05 |
| Unknown | 299.2593 | 5.815166 | 0.86 | (0.72, 1.01) | 0.05 |
| Hydroxyhexadecanoic acid isomer 1 | 353.2313 | 4.621917 | 0.84 | (0.66, 1.01) | 0.05 |
| **D: ILA** |  |  |  |  |  |
| **Putative Metabolite ID** | **m/z** | **Retention time, min** | **OR** | **95% CI** | **P** |
| Unknown | 299.1866 | 3.1635 | 0.85 | (0.72, 0.97) | 0.01 |
| Unknown | 299.2594 | 5.562333 | 1.16 | (1.05, 1.28) | 0.01 |
| Unknown | 299.2593 | 5.630167 | 1.14 | (1.01, 1.27) | 0.05 |

Significance defined as $p < 0.05$ . Putative metabolite IDs shown for metabolites with known confirmed identities.
$\beta$ : coefficient of association; 6-HOME: 6-hydroxyoctadecenoic acid; 11-HOME: 11-hydroxy-9-octadecenoic acid; HETrE: hydroxyeicosatrienoic acid; LTB3: leukotriene B3; m/z: mass-to-charge ratio; OR: odds ratio.

## DISCUSSION

We performed molecular profiling of eicosanoids and eicosanoid-related metabolites to evaluate associations with lung function across two community-based samples. We identified 51 metabolites associated with spirometry measures, including 27 associated with obstructive and 14 with restrictive physiology. This included LA derivatives (9-HODE) and LCFAs including palmitic and stearic acid derivatives (hydroxyhexadecanoic and hydroxyoctadecanoic acids), which correlated with lower PPFEV_1_ and FEV_1_/FVC, and higher odds of obstructive physiology across both samples. Fewer metabolites were associated with restrictive physiology, including an LTB3 analog, associated with higher PPFEV_1_, lower odds of restriction, and lower % emphysema. These findings highlight specific bioactive lipids associated with early changes in lung function.

### Associations with Obstruchtive Lung Physiology

The majority of metabolites identified were associated with obstructive physiology, with expected overlapping associations with PPFEV_1_ and FEV_1_/FVC. Many correlated with higher odds of obstruction, including 9-HODE, a pro-inflammatory LA derivative previously implicated in lung injury and airway inflammation. In studies of porcine models with surfactant depletion, lung injury induced through hyperinflation was shown to elevate levels of oxylipins including 9-HODE.(29) Similarly, in hospitalized COVID-19 patients, exhaled breath condensate samples showed significantly higher levels of eicosanoid compounds including 9-HODE, proposing a possible mechanism for the lung injury and respiratory tract damage seen in severe COVID-19 cases.(30) In vitro models of human and bovine polymorphonuclear leukocytes have shown a chemotactic response to 9-HODE, supporting its role in recruiting pro-inflammatory mediators into airways.(31) We expand upon these findings and show robust association of 9-HODE with obstructive lung physiology, partly mediated by smoking status, consistent with the hypothesis that inhalation insults contribute to activation of inflammatory pathways that impair lung function.

Similarly, we found two LCFAs associated with obstructive physiology. Hydroxyoctadecenoic acid, an unsaturated fatty acid, and isomers of its derivative hydroxyoctadecanoic acid correlated with lower FEV_1_/FVC and higher odds of obstructive physiology in both MESA and FHS. As a naturally-occurring peroxisome proliferator-activated receptor γ (PPARγ) ligand, hydroxyoctadecanoic acid inhibits surfactant protein B gene expression in the lung, disrupting surfactant homeostasis.(32) While patients with chronic asthma are thought to have normal baseline surfactant, in exacerbations surfactant homeostasis disruption is thought to contribute to symptoms. Cigarette smoking and COPD also lead to surfactant dysregulation, contributing to lung function decline.(33) Lastly, isomers of a separate LCFA, hydroxyhexadecanoic acid, also correlated with lower FEV_1_/FVC and obstructive physiology in both MESA and FHS, although its associations with lung function remain unexplored and requires future investigation.

### Associations with Restrictive Lung Physiology

While we found 14 metabolites associated with restrictive physiology in MESA, only three that were associated with lower odds validated in FHS. All three were novel putative eicosanoids with unknown exact molecular identity. Within MESA, we found that EPA, an omega-3 polyunsaturated fatty acid, was associated with higher odds of restrictive ventilatory deficit in MESA and FHS. EPA, converted from alpha linolenic acid or obtained via dietary intake, is anti-inflammatory via inhibition of AA and function as precursors to pro-resolving mediators.(34) Omega-3 fatty acids have been shown to mitigate injury from cigarette smoke induced lung inflammation, bleomycin induced pulmonary fibrosis in mice, and mouse models of acute respiratory distress syndrome.(35–37) Additionally, in a prior study of participants in MESA with interstitial lung disease (ILD), circulating higher levels omega-3 fatty acids were associated with a lower risk of adverse outcomes as well as in healthy MESA participants a slower rate of lung function decline.(38,39) Although it is not clear why we found an association between elevated levels of these omega-3 fatty acids and increased odds of restrictive physiology, it is possible that elevated levels of these anti-inflammatory mediators may have disease mitigating and possibly protective effects in this ostensibly healthy population with presumed early stage disease. However, further research is needed regarding the specific mechanism underlying ILD and the role of elevated levels of omega-3 fatty acids.

### Associations with Lung Structure

We found consistent associations of leukotrienes with CT-based lung imaging measures. Specifically, LTB3 and its analog were associated with greater odds of HAA, a marker of subclinical ILD. A pro-inflammatory AA derivative, LTB3 has not yet been implicated in lung disease, however LTB4, with equipotent pro-inflammatory effects, is involved in neutrophilic pulmonary inflammation and murine emphysema pathogenesis.(40) Interestingly, HAA are associated with other inflammatory biomarkers including matrix metalloproteinase-7 and IL-6,(25) and supporting the role of LTB3 as a potential contributor.

### Limitations

Our study has limitations worth noting. As an observational cross-sectional study, we cannot infer causal relationships between eicosanoid pathways and lung diseases. Longitudinal sampling, especially in response to anti-inflammatory medications, would enable causal inference. Second, lipidomic profiling in MESA was performed one exam cycle before spirometry, introducing potential temporal and selection biases. Further, some eicosanoids were novel molecules with undetermined identities, necessitating future chemistry-based studies.

Diffusion capacity was unavailable in MESA, limiting inferences based on DLCO in this sample, though complementary lung imaging was assessed. FHS participants were predominantly white, limiting generalizability of the validation sample, though MESA was more diverse.

### Summary

Across two large community-based samples of ostensibly healthy adults, we identified 51 eicosanoid and eicosanoid-related metabolites associated with lung structure and function. Pro-inflammatory LA derivatives and LCFAs affecting cellular signaling and surfactant production were associated with obstructive physiology. Pro-inflammatory LTB3 and its analog were associated with emphysema and HAA on imaging. These findings highlight bioactive lipid pathways that may influence lung disease pathogenesis and inform future therapeutic strategies.

## Data Availability

All data produced in the present study are available upon reasonable request to the authors.

## ACKNOWLEDGEMENTS

The authors thank the other investigators, the staff, and the participants of the MESA study for their valuable contributions. A full list of participating MESA investigators and institutions can be found at http://www.mesanhlbi.org.

