## Supplementary Figure 1 for "Association of Eicosanoids and Lung Function in MESA Lung and Framingham Heart Studies"

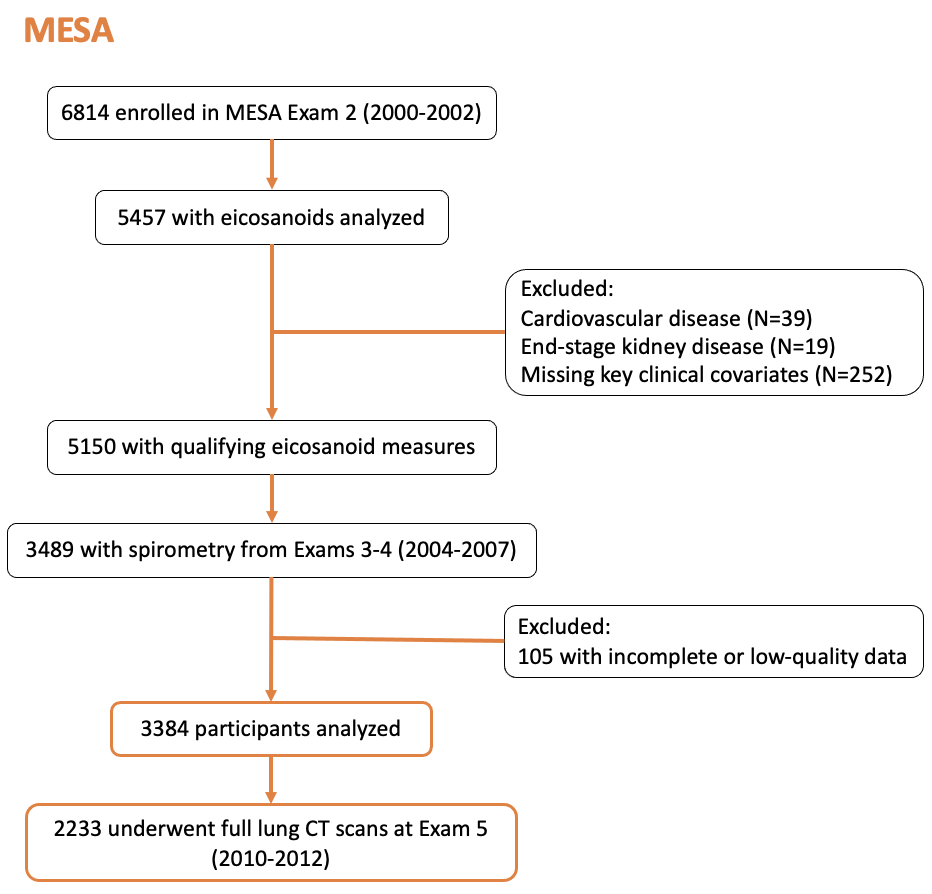

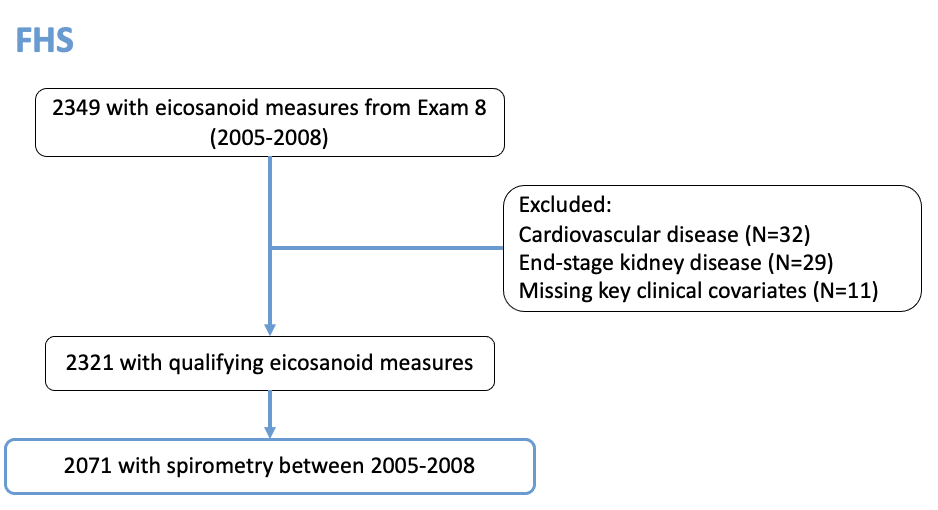

**Supplementary Figure 1:** Timing of data collection and sample selection criteria for MESA and FHS cohorts. Final samples used for analysis shown in orange (MESA) and blue (FHS).
