## Supplementary Figure 2 for "Association of Eicosanoids and Lung Function in MESA Lung and Framingham Heart Studies"

| **A)**  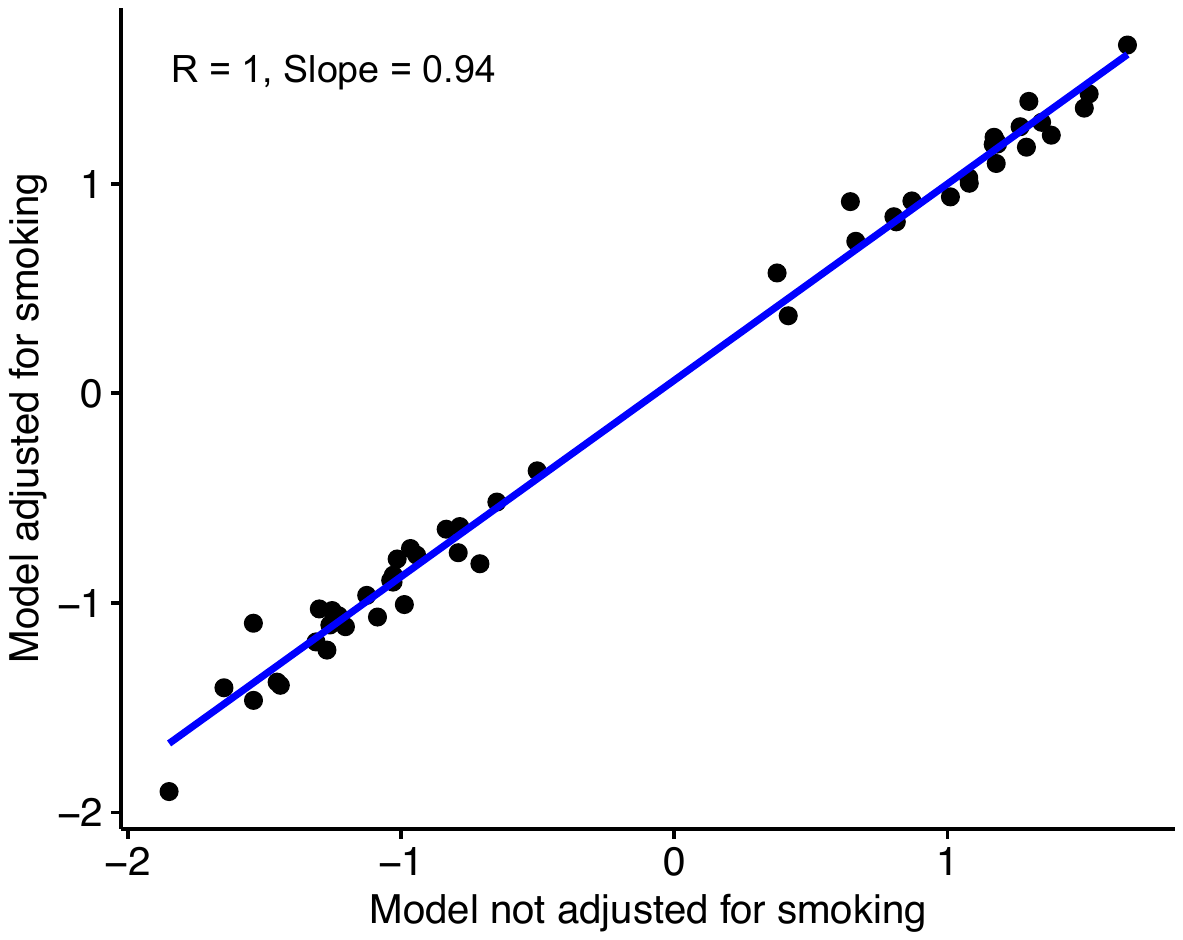 | **B)**  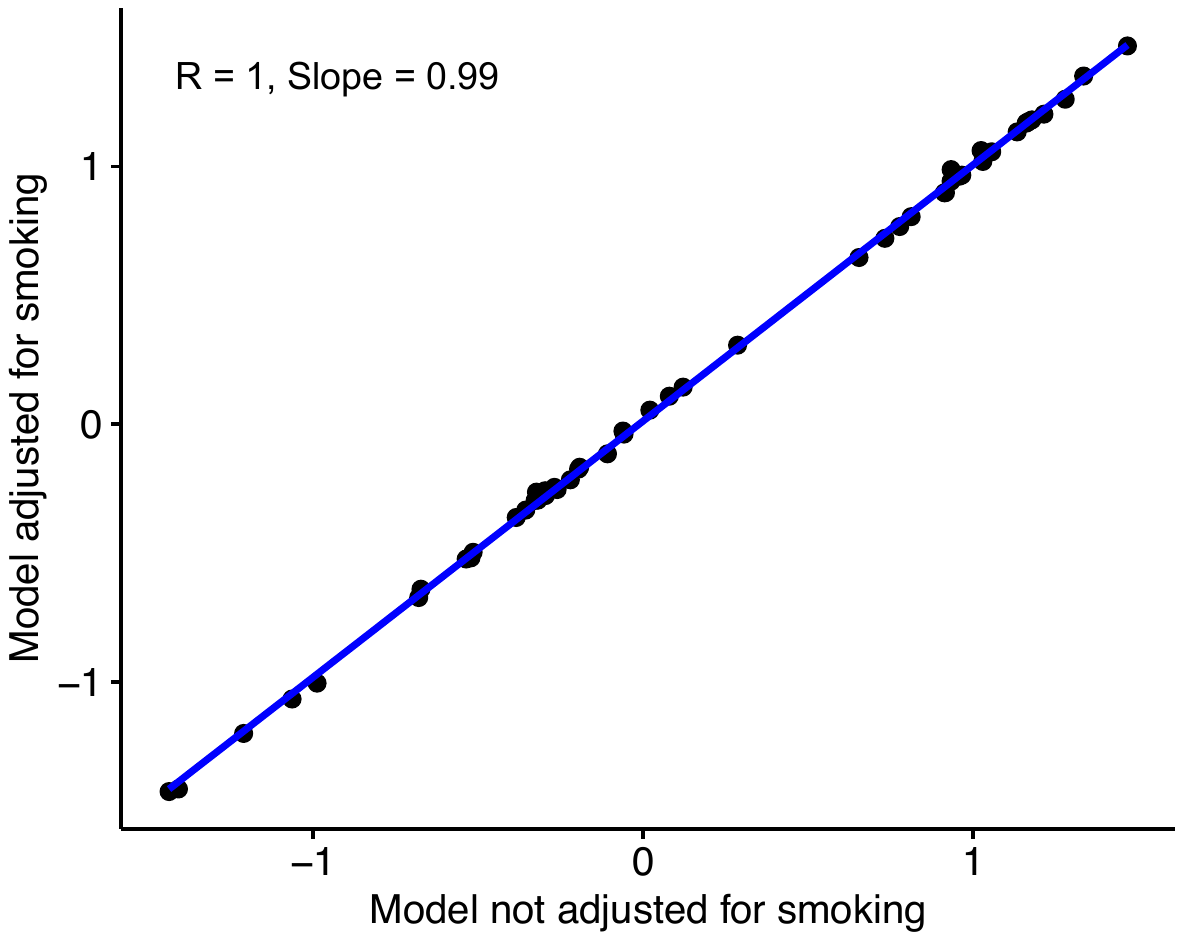 |
| --- | --- |
| **C)**  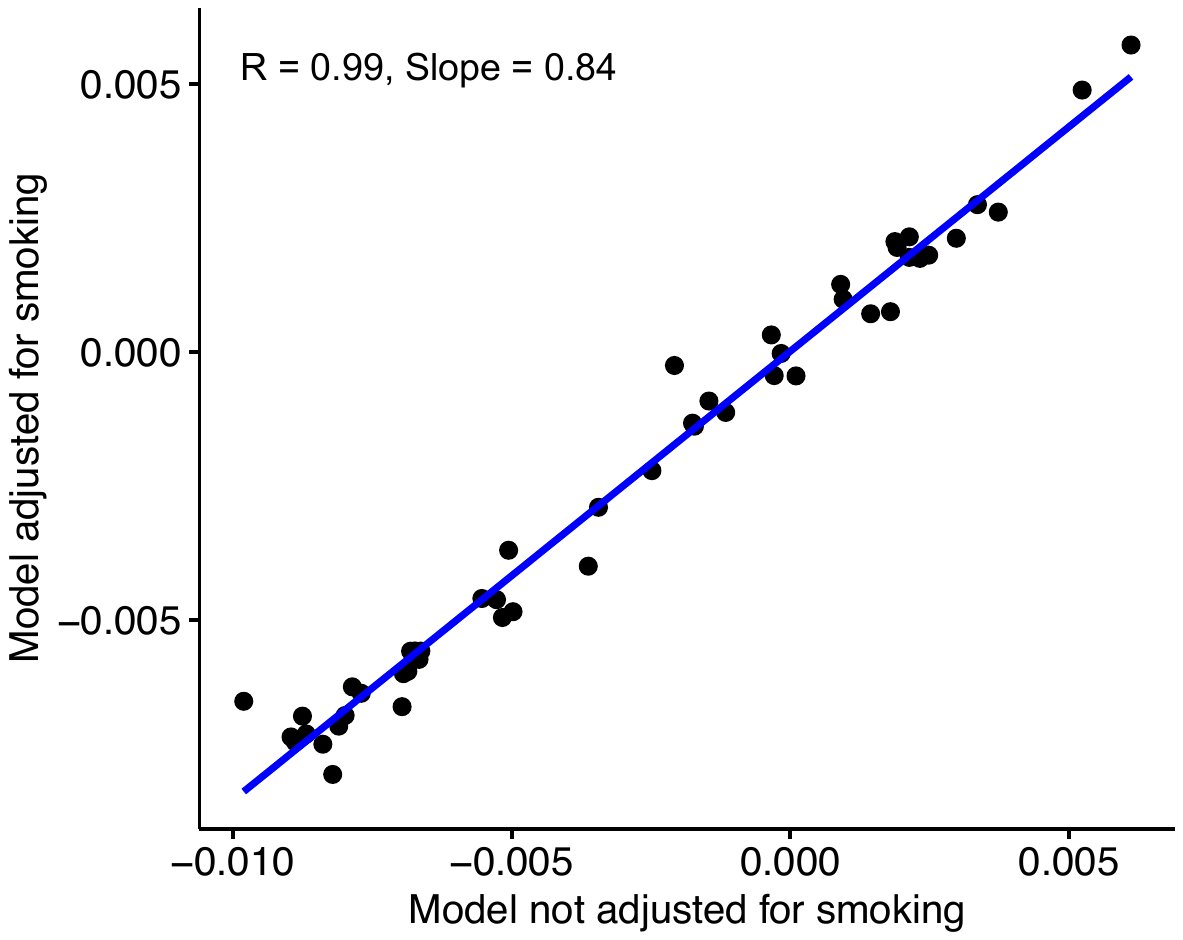 | |

**Supplementary Figure 2:** Relationships between beta coefficients of primary analysis multivariable linear regression model with beta coefficients of models additionally adjusted for smoking status for PPFEV_1_ (A), PPFVC (B), and FEV_1_/FVC (C) models. In general, effect sizes are slightly reduced after adjusting for smoking status.
