## Supplementary Figure 3 for "Association of Eicosanoids and Lung Function in MESA Lung and Framingham Heart Studies"

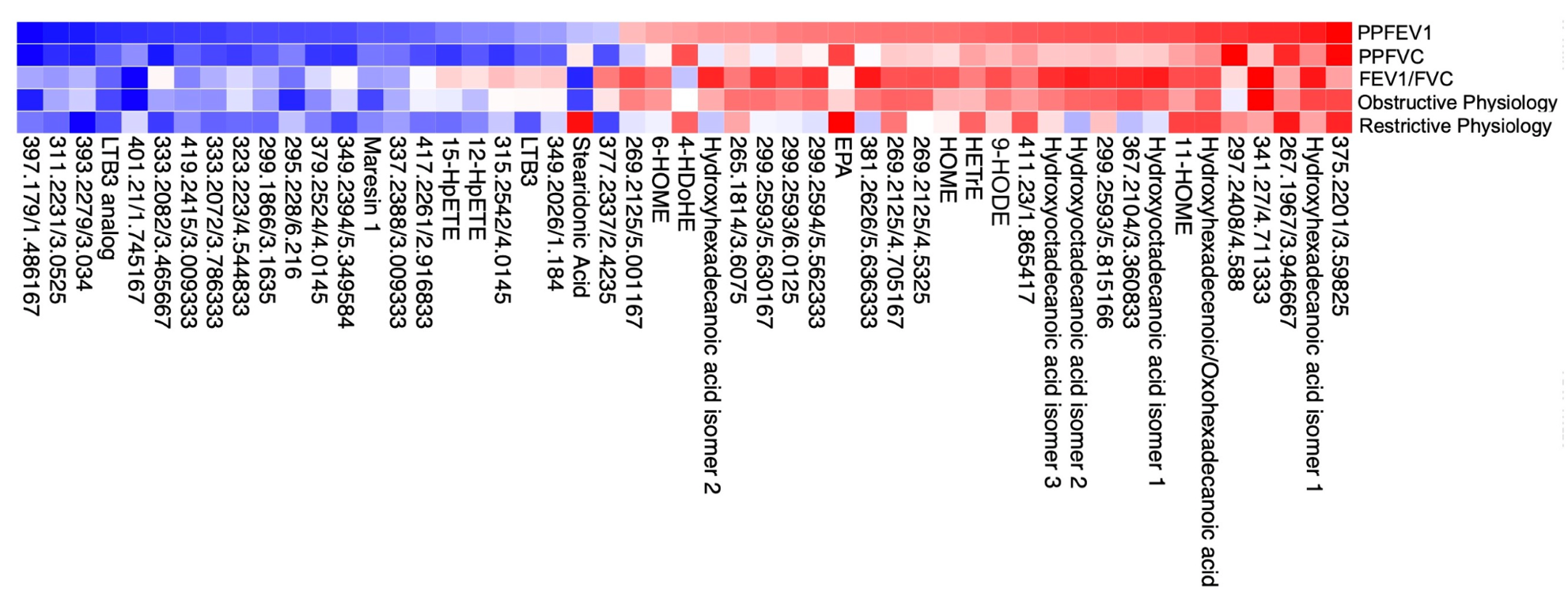

**Supplementary Figure 3:** Heatmap illustrating directionality for associations of eicosanoids and related metabolites with lung function traits, including PPFEV_1_, PPFVC, FEV_1_/FVC ratio, obstructive physiology, and restrictive physiology. Eicosanoid metabolite are annotated by name or mass to charge ratio and retention time. Color scale represents beta coefficients for continuous traits and odds ratios for obstructive and restrictive physiology, scaled to maximum and minimum values, with red indicating worse lung function and blue indicating better lung function (heatmap created using Morpheus).
